# Economic evaluation of a wearable-based intervention to increase physical activity among insufficiently active middle-aged adults

**DOI:** 10.1101/2024.06.05.24306788

**Authors:** Jack H. Ching, Steve Duff, John Hernandez

## Abstract

**Background:** Physical activity levels worldwide have declined over recent decades, with the average number of daily steps decreasing steadily since 1995. Given that physical inactivity is a major modifiable risk factor for chronic disease and mortality, increasing the level of physical activity is a clear opportunity to improve population health on a broad scale. The current study aims to assess the cost-effectiveness and budget impact of a Fitbit-based intervention among healthy, but insufficiently active, adults to quantify the potential clinical and economic value for a commercially insured population in the U.S.

**Methods:** An economic model was developed to compare physical activity levels, health outcomes, costs, and quality-adjusted life-years (QALYs) associated with usual care and a Fitbit-based intervention that consists of a consumer wearable device alongside goal setting and feedback features provided in a companion software application. Improvement in physical activity was measured in terms of mean daily step count. The effects of increased daily step count were characterized as reduced short-term healthcare costs and decreased incidence of chronic diseases with corresponding improvement in health utility and reduced disease costs. Published literature, standardized costing resources, and data from a National Institutes of Health-funded research program were utilized. Cost-effectiveness and budget impact analyses were performed for a hypothetical cohort of middle-aged adults.

**Results:** The base case cost-effectiveness results found the Fitbit intervention to be dominant (less costly and more effective) compared to usual care. Discounted 15-year incremental costs and QALYs were -$1,257 and 0.011, respectively. In probabilistic analyses, the Fitbit intervention was dominant in 93% of simulations and either dominant or cost-effective (defined as less than $150,000/QALY gained) in 99.4% of simulations. For budget impact analyses conducted from the perspective of a U.S. Commercial payer, the Fitbit intervention was estimated to save approximately $6.5 million dollars over 2 years and $8.5 million dollars over 5 years for a cohort of 8,000 participants. Although the economic analysis results were very robust, the short-term healthcare cost savings were the most uncertain in this population and warrant further research.

**Conclusions:** There is abundant evidence documenting the benefits of wearable activity trackers when used to increase physical activity as measured by daily step counts. Our research provides additional health economic evidence supporting implementation of wearable-based interventions to improve population health, and offers compelling support for payers to consider including wearable-based physical activity interventions as part of a comprehensive portfolio of preventive health offerings for their insured populations.

## Introduction

Physical activity (PA) levels worldwide have declined over recent decades, with the average number of daily steps decreasing steadily since 1995.^1^ Among those in high-income countries, nearly 37% do not meet recommended levels of physical activity, and the rates of physical inactivity increase with age.^2^ Inadequate physical activity is associated with substantial healthcare costs and higher risk of incident diseases, such as cardiovascular disease, type 2 diabetes, cancer, and mental health conditions. If current prevalence remains unchanged, physical inactivity is projected to result in 500 million new cases of preventable disease globally, which will incur an additional $300 billion in treatment costs over 10 years.^3^ In the United States (U.S.), less than 50% of the overall population meet the recommended aerobic physical activity guidelines of at least 150 minutes of moderate-intensity exercise per week. This percentage is even lower among older age groups, with only 18-30% of adults ages 35-64 and 10-15% of adults older than 65 meeting these guidelines in 2020.^4^

Given that physical inactivity is a major modifiable risk factor for chronic disease and mortality, increasing the level of physical activity is a clear opportunity to improve population health on a broad scale. There is overwhelming evidence for the health benefits of increased physical activity, including reductions in all-cause mortality, improved cardiorespiratory, metabolic, and muscular health, and reductions in the risk of developing breast and colon cancer.^5–8^ Studies also support the role of regular physical activity in improving sleep quality and mental health outcomes such as anxiety and depression.^9–12^ Recent evidence from the All of Us Research Program, the largest longitudinal study with objectively measured physical activity in the U.S., re-affirmed these benefits among participants who used a Fitbit device to track their physical activity. Those who maintained higher daily step counts over a median follow-up of 4 years had 25-46% lower risks of developing multiple chronic conditions, including diabetes, hypertension, obesity, sleep apnea, gastroesophageal reflux disease, and major depressive disorder.^13,14^

Wearable activity trackers, such as Fitbit devices, offer a promising low-cost approach to help increase physical activity levels, leading to substantial health benefits. These trackers can be used either as the primary component of an intervention (often in conjunction with a software application) to promote self-monitoring and goal setting, or as a part of more comprehensive programs that also incorporate additional counseling, education, and/or structured exercise elements.^15,16^ Across more than a hundred randomized controlled trials (RCTs), these interventions consistently show clinically significant increases in daily step counts, increases in moderate to vigorous physical activity (MVPA), and improvements in body composition, blood pressure, and fitness.^16,17^ Among healthy adults, wearable-based interventions were associated with improvements in physical activity over an average of 21 weeks, and long-term follow-up found that approximately 40% of the short-term improvements were maintained through 3-4 years.^18,19^ These improvements are larger among older adults and those with chronic conditions such as type 2 diabetes and COPD, for whom wearable activity interventions resulted in significant short-term improvements in physical activity and reduced sedentary time.^20,21^ It is also notable that even the provision of a pedometer alone, without additional human-delivered behavioral change interventions, can increase both daily steps and MVPA minutes for up to 12 months.^22^

Fitbit devices, in particular, have been effective in promoting healthy lifestyle behaviors and supporting chronic condition management as part of multifaceted behavior change interventions. A systematic review of 37 RCTs found that Fitbit-based interventions increased the number of daily steps, minutes of weekly activity, and led to additional pounds of weight lost.^23^ The evidence for Fitbit-based interventions in sub-populations are consistent with those for wearable activity trackers in general. Studies among sedentary adult women found that the Fitbit-based intervention substantially increased steps per day, and the number of bouts and minutes of MVPA per week.^24,25^ Fitbit-based interventions have also been shown to increase physical activity levels in older adults for up to 32 weeks, and to increase various measures of physical fitness in this population.^26–28^

There is also abundant support for the health economic value of greater physical activity over the long-term, and for wearable-based interventions that aim to increase physical activity levels. Adopting or maintaining a physically active lifestyle throughout adulthood is associated with reductions in Medicare costs later in life, compared to those who remained inactive.^29^ Wearable-based physical activity interventions, ranging from those that use pedometers, exercise prescriptions, individual/group counseling, to mass media promotions, have all been shown to be cost-effective or cost-saving among populations of middle-aged and older adults.^30–34^ In general, studies have found less costly interventions, such as those focused on self management using pedometers, to be more cost-effective or more likely to be cost-saving when compared to higher cost interventions that also include physician visits and/or referral to exercise professionals.

Building on existing health and economic evidence for wearable-based interventions that focus on improving physical activity, the current study aims to assess the cost-effectiveness and budget impact of a Fitbit-based intervention (consisting of a consumer wearable device and app-based goal setting and feedback features) among healthy, but Economic Evaluation of a Wearable-based Intervention insufficiently active, adults to quantify the potential clinical and economic value for a commercially insured population in the U.S.

## Methods

### Modeling Approach

A Markov cohort state-transition model was developed to quantify the clinical, economic, and humanistic outcomes of usual care and a Fitbit intervention aimed at increasing physical activity in adults who are insufficiently active. The modeling approach was informed by previous research^30–39^ and good research practices published by the International Society for Pharmacoeconomics and Outcomes Research (ISPOR).^40–45^ A schematic in **Figure 1** illustrates the modeling framework, disease health states, and outcomes.

**Figure 1.**
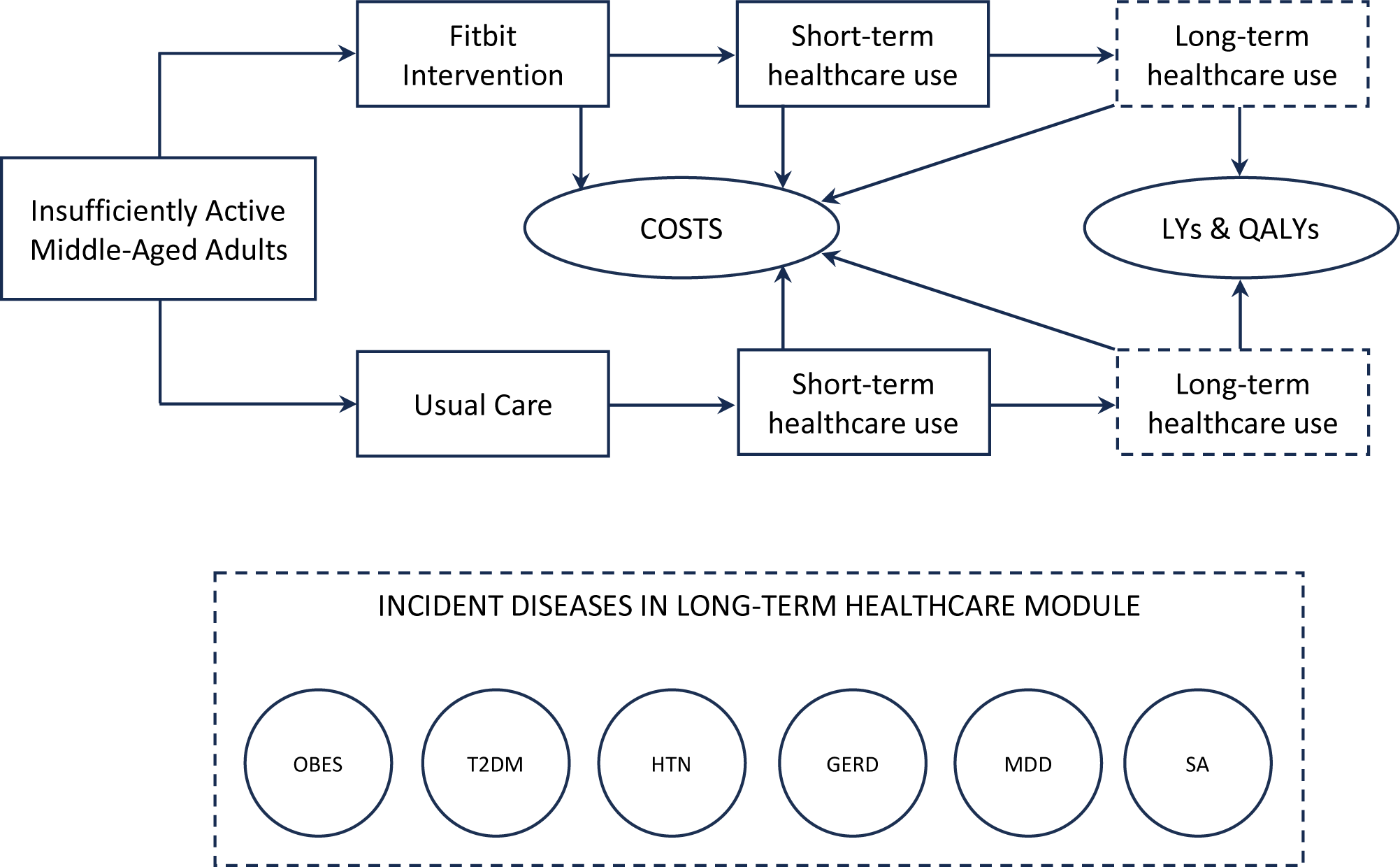
A simplified schematic of the physical activity intervention model. During and after a physical activity intervention (Fitbit or Usual Care), short-term (years 1 and 2) and long-term (post-2 years) healthcare resource use and costs are quantified. The long-term impact of the interventions on 6 conditions associated with insufficient physical activity also is considered in terms of survival and quality-adjusted survival. Abbreviations: GERD, gastroesophageal reflux disease; HTN, hypertension; LY, life-year; MDD, major depressive disorder; OBES, obesity; QALY, quality-adjusted life-year; SA, sleep apnea; T2DM, type 2 diabetes mellitus

The model was designed to estimate the impact of the Fitbit intervention on physical activity in terms of an increase in the mean number of daily steps. In addition to being readily accessible and widely used to measure physical activity, daily step count is a well-supported metric that is inversely and linearly associated with health outcomes including cardiovascular disease events, type 2 diabetes incidence, and all-cause mortality.^46^ In addition, total daily step counts and other physical activity measures have been found to have an effect on both short-term (i.e., 1-2 years) medical resource utilization and costs^47–52^ while the impact on longer-term costs and quality of life has been quantified in numerous modeling studies through decreased incidence of diseases associated with sedentary behavior.^30–39^ The cohort of individuals in the analysis were assumed to be in general good health at baseline (i.e., devoid of any of the chronic diseases of interest) and started in the ‘Well’ health state. During each model cycle (1 year), individuals remain in the ‘Well’ health state or transition to any of the incident disease health states or death, based on background age/sex-based mortality, adjusted according to health state-specific mortality hazard ratios. Transitions into one of the chronic disease states precluded transition back to the ‘Well’ health state; participants only remain in the disease health state or transition to the death absorbing state. Costs, life-years (LYs) and quality-adjusted life-years (QALYs) are accrued in each cycle.

### Population

The target population of interest for the intervention in the analysis was insufficiently active adults in the U.S., aged 40-60 (mean age 50) who are insured by Commercial health plans. The age and sex distribution was based on 2022 data from the U.S. Census Bureau.^53^ The proportion of adults with insufficient physical activity in this population was informed by a recent nationwide survey study of 2,640 participants.^54^ Population details are shown in **Table 1**.

**Table 1.**
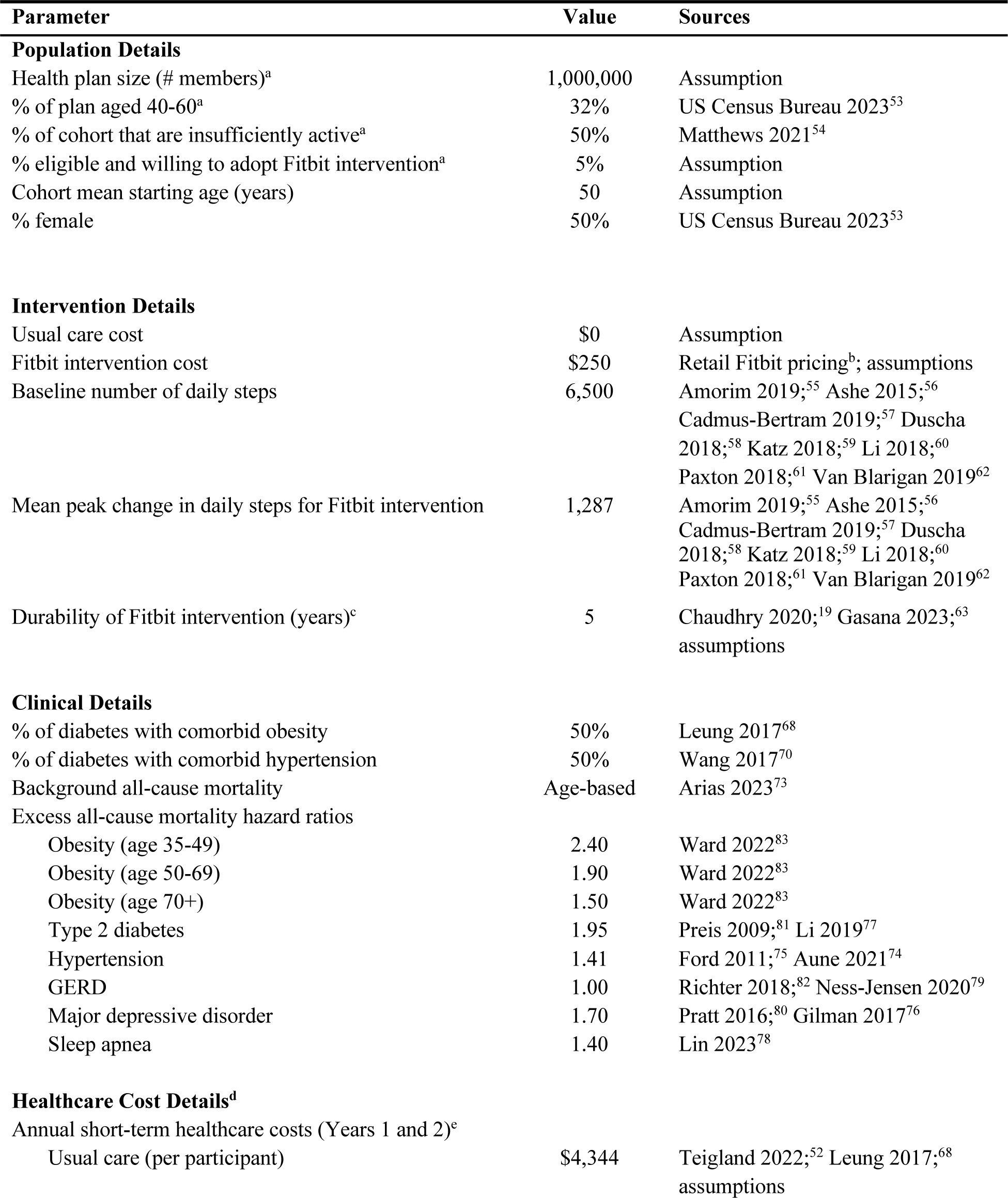
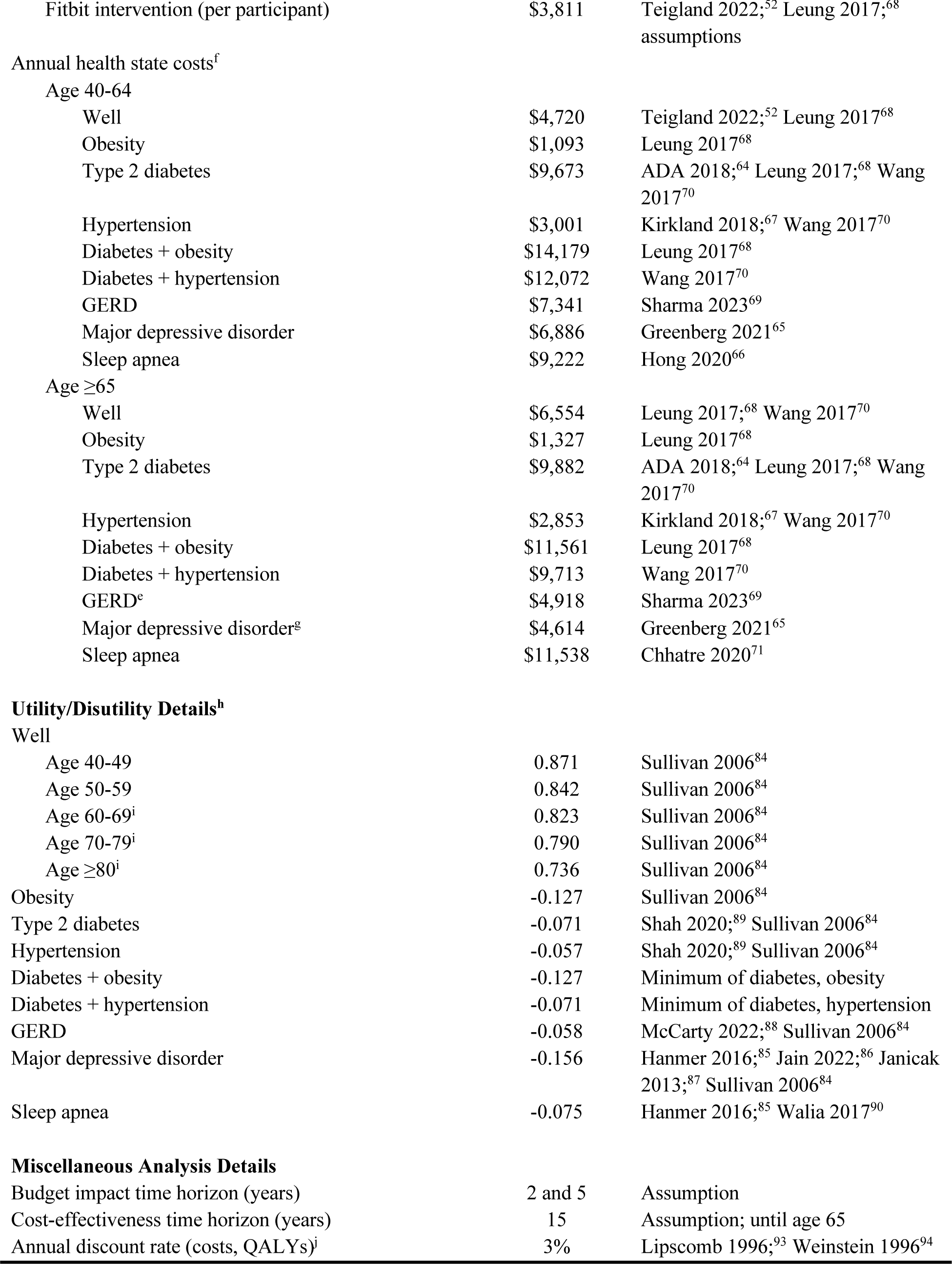
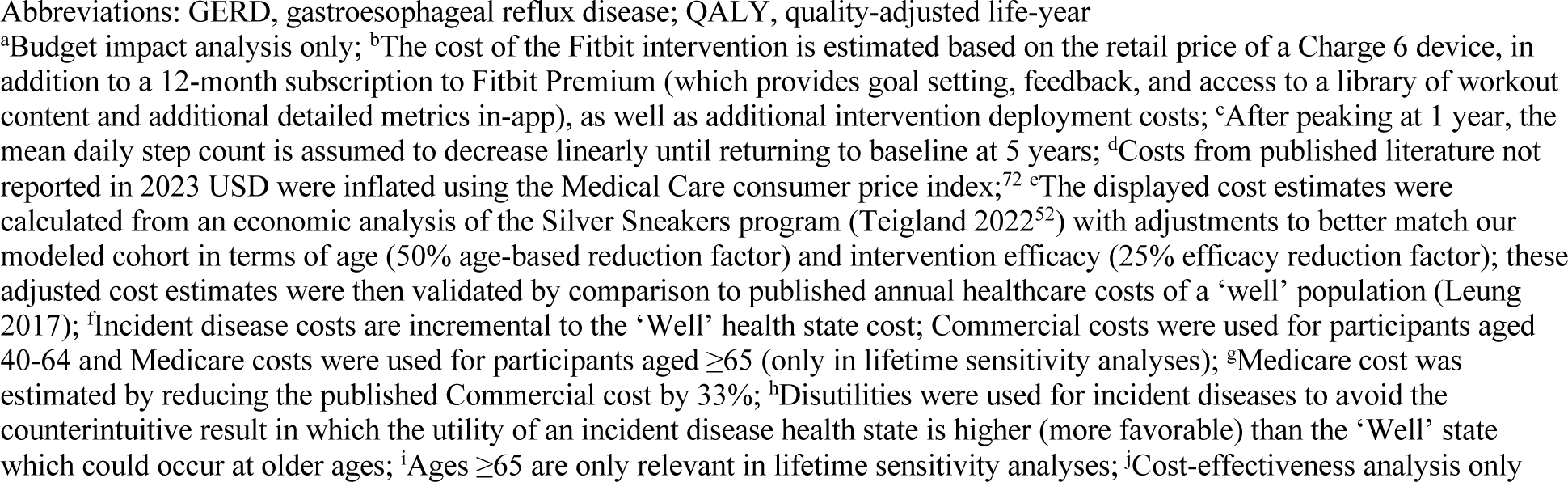
Base Case Model Parameter Estimates and Sources.

### Comparators

As described above, physical activity interventions using wearable activity trackers such as accelerometers and pedometers have resulted in improved physical activity and clinical outcomes. A common alternative assessed in evaluations of physical activity interventions is usual care, which is the comparator we adopted for our economic analysis. Unlike usual care, which does not specifically encourage or increase physical activity and therefore results in no meaningful or lasting change in daily step count, the Fitbit intervention is estimated to increase the average daily step count by 1,287 steps, or approximately 20% from a baseline daily step count of 6,500. These estimates were derived using data from 8 RCTs of Fitbit-based interventions among participants less than 65 years of age (**Supplementary Text**).^55–62^ The durability of the Fitbit intervention was informed by two systematic reviews and meta-analyses of 8 and 57 RCTs (8,480 and 16,355 participants, respectively) that determined step count monitoring interventions to have sustained benefits over at least 4 years.^19,63^ Both analyses demonstrated statistically significant increases in daily step counts in the intervention group over controls, although the magnitude of the increases waned over time. To emulate this in the model, the maximum step count increase for the Fitbit intervention was assumed to peak at 1 year, decline linearly by 25% per year, with average daily step counts returning to baseline (and equivalent to usual care step count levels) beginning at year 5 (**Table 2**).

**Table 2.** Intervention Efficacy (Mean Daily Steps per Day)^a^.

| Mean Number of Daily Steps | Year 0 | Year 1 | Year 2 | Year 3 | Year 4 | Year 5 | Year 6 | Year 7<sup>b</sup> |
| --- | --- | --- | --- | --- | --- | --- | --- | --- |
| Usual Care Cohort | 6,500 | 6,500 | 6,500 | 6,500 | 6,500 | 6,500 | 6,500 | 6,500 |
| Fitbit Cohort | 6,500 | 7,787 | 7,465 | 7,144 | 6,822 | Equivalent to Usual Care |  |  |
| Incremental Steps | 0 | 1,287 | 965 | 644 | 322 | 0 | 0 | 0 |

Sources: STEPS—Amorim 2019;<sup>55</sup> Ashe 2015;<sup>56</sup> Cadmus-Bertram 2019;<sup>57</sup> Duscha 2018;<sup>58</sup> Katz 2018;<sup>59</sup> Li 2018;<sup>60</sup> Paxton 2018;<sup>61</sup> Van Blarigan 2019;<sup>62</sup> DURABILITY—Chaudhry 2020;<sup>19</sup> Gasana 2023;<sup>63</sup> assumptions
<sup>a</sup>Sensitivity analyses allowed the mean number of daily steps in the Fitbit cohort to return to baseline as early as Year 2 or as late as Year 6; <sup>b</sup>Year 7 daily steps are carried forward in subsequent years

As is common with comparable wearable-based physical activity interventions, the Fitbit intervention was assumed to consist of a consumer wearable device alongside goal setting and feedback features provided in a companion software application. We further assume that the Fitbit intervention does not include additional human-delivered intervention components, and conducted extensive sensitivity analyses to explore our assumptions of intervention effectiveness.

To model the incidence of chronic diseases related to insufficient physical activity—including obesity (OBES), type 2 diabetes mellitus (T2DM), hypertension (HTN), gastroesophageal reflux disease (GERD), major depressive disorder (MDD), and sleep apnea (SA)—we utilized new analyses of previously published estimates of disease incidence derived from electronic health records (EHR) in the All of Us Research Program.^13^ This unique ongoing research program is funded by the National Institutes of Health and is currently collecting multiple streams of health-related information from hundreds of thousands of Americans. As of the 2022 publication by Master et al., 6,042 of the 214,206 program participants sharing their EHR data had linked their own Fitbit devices to the data collection effort, thereby providing a rich data source on the relationship between physical activity and the risk of developing chronic disease. Cumulative incidence estimates by year (3, 5, and 7) for six diseases based on a linear function (as opposed to the published estimates based on a cubic spline fit) of average daily step count were obtained from the All of Us researchers for model parameterization to mitigate potential overfitting. Cumulative incidence data were converted to the annual incidence rates as model inputs (**Supplementary Table 1**).

### Costs

The costs in the model include those related to the Fitbit intervention (incremental to usual care which is assumed to have no intervention cost), overall short-term healthcare costs in Years 1 and 2, and longer-term health state-related costs (based on ‘Well’ and disease states) in Years 3+. Given the non-invasive nature of the Fitbit intervention, adverse events are uncommon and assumed to have nominal costs; therefore, they were not included in the analysis. Unit costs for the model were derived primarily from published sources.^52,64–71^ In cases where robust evidence was unavailable, conservative assumptions based on clinical expert opinion were used to supplement published estimates. Furthermore, in several instances, published costs required conversion between the Medicare and Commercial perspectives. Inflation and deflation factors of 50% (Medicare to Commercial) and 33% (Commercial to Medicare), respectively, were utilized for this purpose. All unit costs in the model are reported in 2023 U.S. dollars (USD); published costs were inflated using the Medical Care component of the U.S. consumer price index (CPI)^72^ when necessary (**Table 1**).

The cost of the Fitbit intervention is estimated based on the retail price of a Charge 6 device, in addition to a 12-month subscription to Fitbit Premium (which provides goal setting, feedback, and access to a library of workout content and additional detailed metrics in-app), as well as additional intervention deployment costs.

Numerous studies have documented the impact of increased physical activity (using various modalities or interventions) on short-term medical resource use and costs, usually measured over 1-2 years.^47–52^ Given the substantial and clinically meaningful increase in average daily steps observed with Fitbit use,^63^ we assumed a potential for modest short-term reduction in resource use and costs in our analyses. This model element was informed by an analysis of claims data from over 8,500 Medicare Advantage enrollees participating in the Silver Sneakers*^Q^*^R^ (SS) healthy aging program.^52^ Because the study population and intervention are not an exact match to those in our model, two factors were included in our model to reduce the potential cost savings due to differences in population age (reflecting an assumption of lower overall healthcare costs in a younger population) and intervention intensity (reflecting an assumption that the SS program has a greater impact on resource use/cost). These parameters and conservative assumptions were evaluated extensively in both one-way and probabilistic sensitivity analyses as well as with comparison to published estimates of annual healthcare costs in a comparably aged population.^68^

Finally, longer-term costs associated with the model health states (‘Well’ and incident diseases) were quantified for both comparators. Incremental costs attributable to each disease health state were extracted from the literature and added to the ‘Well’ health state to yield costs for each of the incident disease states.^52,64–71^

### Mortality

The annual probability of death for participants in the ‘Well’ health state was based on age- and sex-adjusted all-cause mortality data from U.S. life tables.^73^ Published hazard ratios describing the excess mortality associated with each of the chronic diseases of interest were used to adjust background all-cause mortality and the annual probability of death for those disease states.^74–83^

### Utilities

Health-related quality of life utility and disutility weights were used to calculate QALYs for the cost-effectiveness analysis (CEA). Utilities for the ‘Well’ health state (by age strata) were based on a nationally representative catalog of EuroQol-5 Dimension (EQ-5D) utility scores.^84^ For the chronic diseases in the model, we identified published disutilities^85–90^ (or utilities that were subsequently used to derive disutilities using information in Sullivan, et al. 2006^84^) that were applied to the ‘Well’ health state utility to account for the worse quality of life for those conditions.

### Validation and Analyses

Consistent with best practices guidance, the model was subjected to extensive validity testing.^41,42,44^ The face validity of the model was evaluated by comparison to other physical activity intervention model structures and through a clinical applicability review (J.H.). Internal validity was assessed by a review of the model programming by a second modeler (J.H.C.) and through a series of quality assurance tests and analyses to confirm proper model functionality. Finally, the external validity of the epidemiologic framework was appraised by comparisons of model outcomes to U.S. life expectancy data and other published studies.^73,91,92^

Upon conclusion of the validation process, base case cost-effectiveness and budget impact analyses using the default parameter estimates were conducted. Both types of economic analysis took the perspective of a Commercial payer. Therefore, for the CEA, the time horizon was set to 15 years when the mean age in the model reached 65 (i.e., when a Medicare payer perspective would be more relevant). For the budget impact analysis (BIA), two separate time horizons were evaluated. A 2-year horizon was evaluated to focus on the potential short-term clinical and cost benefits while a 5-year horizon was used to also capture longer-term benefits and to be consistent with the modeled durability of the Fitbit intervention. Outcomes were discounted at an annual rate of 3% in the CEA and were not discounted in the BIA as recommended in good research practice guidelines.^45,93,94^ Although guidelines also recommend conduct of analyses from the societal perspective, the simple, non-invasive nature of the intervention is unlikely to result in substantial impacts within the informal health care and non-health care sectors, such that this would not materially change our conclusions.

The impact of uncertainty in parameter estimates and model structure was evaluated with deterministic and probabilistic sensitivity analyses (PSA). Deterministic analyses included one-way sensitivity analyses (OWSA) and scenarios constructed to assess multi-parameter variability. Probabilistic analyses included over 40 parameters varied simultaneously with new estimates drawn at random from their respective probability distributions (e.g., beta distribution for utilities, gamma distribution for costs, etc.) for 5,000 runs. Net monetary benefit (NMB) was calculated at an assumed willingness to pay (WTP) of $150,000/QALY.^95,96^ Cost-effectiveness acceptability curves and a scatterplot were also constructed based on the results of the PSA.

Finally, an exploratory analysis of an insufficiently active older population (mean starting age 65) was conducted to assess the costs and outcomes of the Fitbit intervention from a Medicare payer perspective. A recent publication by Deidda, et. al.^31^ informed the exploratory analysis. Analysis details and results are presented in the **Supplementary Materials**.

## Results

### Validation Results

The model was deemed valid based on the evaluations conducted and the external validity check. For the latter, we appraised the epidemiologic framework to ensure that our modeling of incident disease and survival resulted in credible predictions. For both the base case population (middle-aged adults; mean age 50) and exploratory population (older adults; mean age 65), our model estimated mean life expectancy 2.5 – 3 years less than that based on U.S. life table predictions.^73^ This is consistent with studies from Australia and the U.S. documenting a life expectancy reduction of 1.5 – 4.8 years due to sedentary behavior^92^ or, conversely, an increased life expectancy of 2 years if sedentary behaviors were to be ameliorated.^91^

### Base Case Results

The results of the base case economic analyses are presented in **Table 3** and **Figure 2**. In the cost-effectiveness analysis, the total 15-year discounted costs of the Fitbit intervention were less than for usual care alone ($88,284 vs $89,541) resulting in a net cost savings of $1,257. The majority of the cost savings ($1,049) was associated with short-term cost reduction with the remainder due to reductions in incident disease. A nominal increase in life-years was observed for the Fitbit intervention (increment of 0.003 LYs) related to lower incident disease mortality. Similarly, because Fitbit use results in a greater proportion of time spent in the ‘Well’ health state, QALYs also are higher than for usual care (9.042 vs 9.031). As the Fitbit intervention is less costly and more effective than usual care, an ICER is not calculated and the Fitbit intervention is considered to “dominate” usual care.

**Figure 2.**
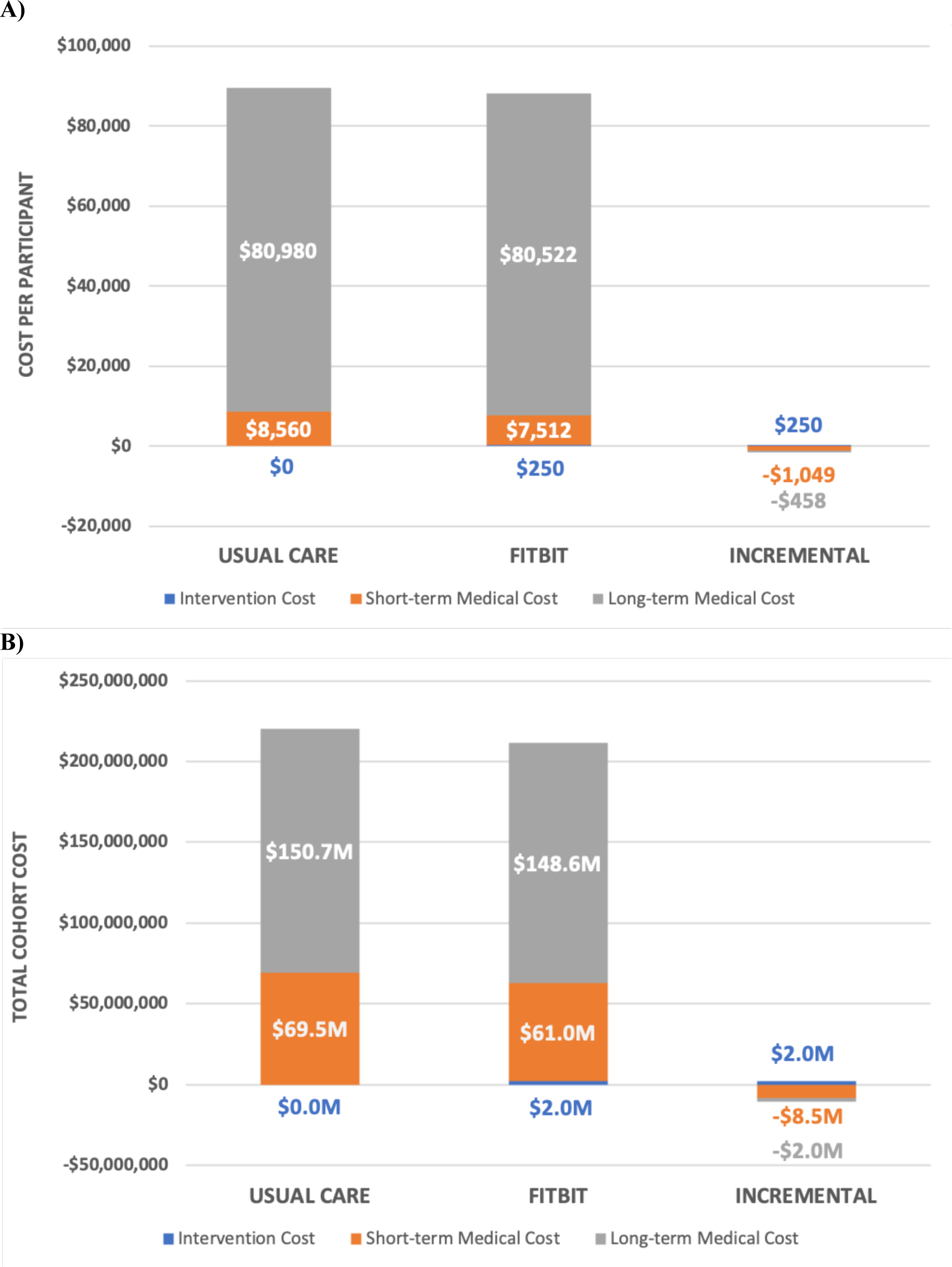
Stratified cost results of the A) cost-effectiveness and B) budget impact analyses. The time horizons of the analyses are 15 years and 5 years, respectively. The majority of the cost savings is associated with reductions in healthcare resource use in the first two years after initiation of the interventions.

**Table 3.** Base Case Economic Analysis Results.

|  | Usual Care | Fitbit | Incremental |
| --- | --- | --- | --- |
| **Cost Effectiveness<sup>a</sup>** |  |  |  |
| Total costs | \$89,541 | \$88,284 | -\$1,257 |
| Total LYs | 11.490 | 11.493 | 0.003 |
| Total QALYs | 9.031 | 9.042 | 0.011 |
| ICER (cost/LY gained) |  |  | Fitbit dominates UC<sup>c</sup> |
| ICER (cost/QALY gained) |  |  | Fitbit dominates UC<sup>c</sup> |
| **Budget Impact<sup>b</sup>** |  |  |  |
| 2-year total costs | \$69,496,000 | \$62,982,000 | -\$6,514,000 |
| 2-year incremental budget impact (PMPM) | | | -\$0.27 |
| 5-year total costs | \$220,166,612 | \$211,617,969 | -\$8,548,644 |
| 5-year incremental budget impact (PMPM) | | | -\$0.14 |

Abbreviations: ICER, incremental cost-effectiveness ratio; LY, life-year; PMPM, per-member per-month; QALY, quality-adjusted life-year; UC, usual care
<sup>a</sup>Expected costs and outcomes per participant quantified over a 15-year time horizon, discounted at 3% annually; <sup>b</sup>Costs for a cohort of 8,000 participants over both a 2-year and 5-year time horizon (undiscounted); <sup>c</sup>“Dominates” indicates that Fitbit is less costly and more effective than UC

Examining the economics from the perspective of a Commercial healthcare payer, in a 1-million member plan, it was estimated that there would be approximately 160,000 members in the 40-60 year old age group that would be classified as having insufficient physical activity. Of those, we assumed that 8,000 would be eligible and choose to adopt a Fitbit intervention and be included in the budget impact analysis.

Total costs for a cohort adopting the Fitbit intervention were lower than if they received usual care and remained insufficiently active (**Table 3**). Two-year and five-year cost savings were $6,514,000 and $8,548,644, respectively which equate to -$0.27 per member per month (PMPM) and -$0.14 PMPM. Given the shorter time horizon in the budget impact analysis, the contribution of incident disease cost savings is even lower than in the cost-effectiveness analysis.

### Sensitivity and Scenario Analysis Results

One-way deterministic sensitivity analyses were conducted by varying key parameter values within credible ranges. Results of the sensitivity analyses are illustrated in a tornado diagram (**Supplementary Figure 1**); however, rather than displaying the results in terms of the impact of parameter changes on the ICER results, the tornado diagram shows the change in net monetary benefit at a willingness to pay threshold of $150,000/QALY since all changes in individual parameters do not result in the calculation of an ICER (i.e., the Fitbit intervention remains dominant). These sensitivity analyses illustrate the importance of the short-term cost parameters on the results with 3 of the top 5 most influential parameters relating to this model element. However, even when these key parameter estimates are varied, the net monetary benefit is positive and the Fitbit intervention remains cost saving (not shown in a Figure). Furthermore, these analyses demonstrate that, even with step count improvements as low as 750 steps per day or if the intervention were only effective for 1 year (i.e., step count levels return to baseline at Year 2), the Fitbit intervention remains dominant.

While the deterministic results are informative regarding individual model parameters, the results of the PSA, in which the uncertainty in numerous parameters is evaluated simultaneously, are much more informative about the robustness of the cost-effectiveness results. The PSA results are presented in the scatterplot in **Figure 3**. Each of the 5,000 model runs is shown as an incremental QALY and cost pair (Fitbit intervention relative to usual care). To facilitate interpretation, lines representing the ICER thresholds of $50,000, $100,000, and $150,000/QALY gained are superimposed on the graph. The vast majority (93.0%) of the simulation results fell into the southeast quadrant of the graph indicating that the Fitbit intervention is dominant (less costly and more effective). Over 99% of the model runs resulted in an ICER less than $150,000/QALY gained or Fitbit intervention dominating usual care. These results provide very high confidence that, even when accounting for parameter uncertainty (including in the key short-term cost parameters), a Fitbit intervention would provide favorable value or cost savings compared to usual care.

**Figure 3.**
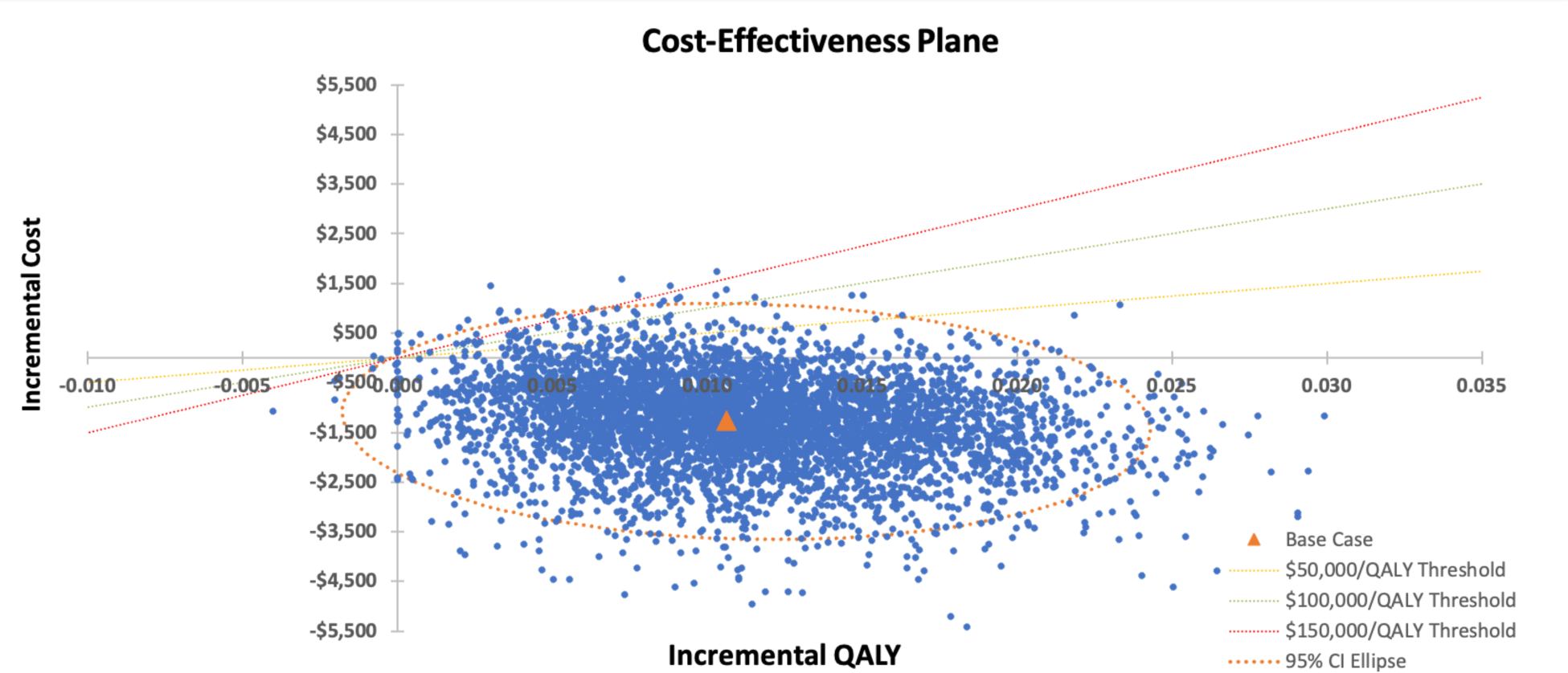
Scatterplot of the probabilistic sensitivity analysis (5,000 model runs). Incremental costs and QALYs of the Fitbit intervention compared to Usual Care are demonstrated. The orange triangle denotes the base case ICER. ICER lines representing $50,000, $100,000, and $150,000/QALY thresholds are displayed as is the 95% confidence interval ellipse. Abbreviations: CI, confidence interval; ICER, incremental cost-effectiveness ratio; QALY, quality-adjusted life-year

**Figure 4.**
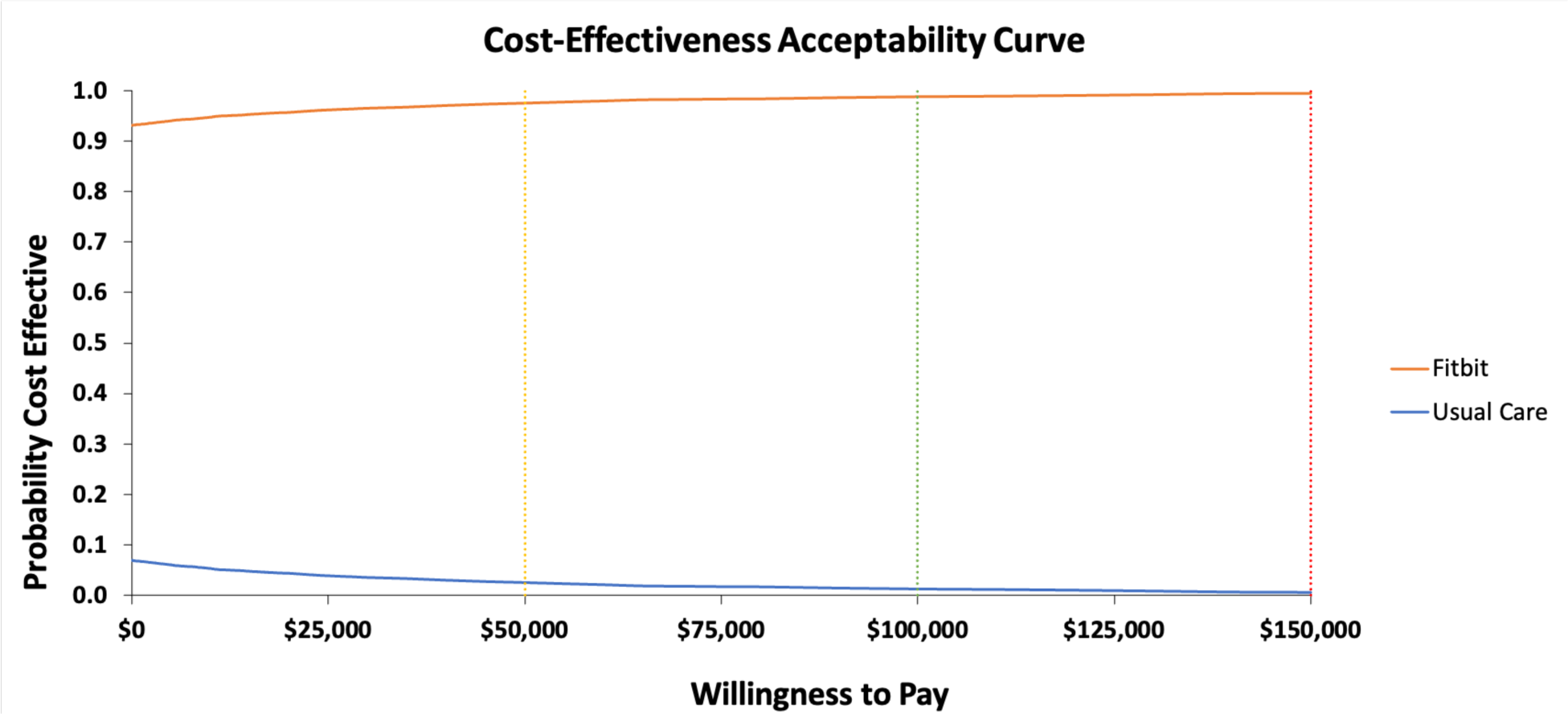
Cost-effectiveness acceptability curve. At all willingness to pay thresholds ($/QALY gained), a physical activity intervention including a Fitbit is the preferred treatment strategy over Usual Care. Abbreviation: QALY, quality-adjusted life-year

Good research practices in modeling call for an examination of structural uncertainty in the model.^41,42,44^ We attempted to address this issue in two ways; first, we evaluated an alternative approach for characterizing intervention effectiveness in terms of daily step counts (a ‘tiered steps approach’ that allows for varying levels of step count increases among population subgroups vs the ‘mean steps approach’ [the model default]) and second, we varied how health state costs were integrated into the analysis (inclusion of these costs in the first 2 years vs. exclusion [the model default]). Modifications to each structural element resulted in a nominal change to the model outcomes, none of which materially changed the interpretation or conclusions of the analysis (**Supplementary Table 3**). More information on the analyses is available in the **Supplementary Materials**.

In addition to the various types of sensitivity analyses described above, we also performed several ad-hoc scenario analyses in which one or more parameters were varied. These included:

- Use of the ‘post-intervention’ steps estimate rather than the ‘difference in difference’ approach described in the **Supplementary Materials**;
- An increase and decrease of the proportion of eligible members adopting a Fitbit intervention (budget impact analysis only);
- Use of the upper and lower bounds of disease incidence estimated by the linear function of daily steps and incidence as obtained from the All of Us researchers;
- Extension of the cost-effectiveness analysis time horizon from 15 years to a lifetime horizon; and
- Substitution of health state costs (‘Well’ and incident diseases) in Years 1 and 2 for the general short-term healthcare cost component.

As presented in **Supplementary Table 3**, modification of the mean daily steps estimate, intervention adoption rates, the cumulative incidence of chronic disease, or the time horizon had a nominal impact on the results with the Fitbit intervention remaining dominant and cost saving compared to usual care. The results changed substantially only when health state costs were the only type of cost considered in the analysis. The Fitbit intervention was dominant and cost saving compared to usual care across the range of scenario analyses with the 15-year per-person cost savings ranging from -$1,257 (when the short-term cost savings were included) to -$318 (when the only source of cost savings was attributable to reduction in the onset of chronic disease). While complete exclusion of the literature-supported overall short-term healthcare costs is an extreme assumption, it illustrates the need for further research to ascertain the potential reduction in short-term healthcare resource use associated with a physical activity intervention in the middle-aged population.

The exploratory analysis of a Fitbit-based intervention in an older population insured by Medicare demonstrated greater cost savings associated with the intervention than in the base case analysis of a middle-aged population (**Supplementary Tables 4 and 5**, **Supplementary Figure 2**). This is primarily due to the magnitude of short-term healthcare cost savings, for which the evidence is more robust in an older population, which is 2-to 4-times greater in the exploratory analysis than in the base case population. As a result, the cost savings associated with incident disease reduction in the exploratory analysis make up a much smaller proportion of the overall cost savings, partially because the inclusion of fewer chronic diseases in the exploratory analysis limits the magnitude of achievable cost savings. In addition, over the lifetime horizon in the exploratory analysis, only a fraction of disease occurrence can be completely prevented by an intervention that, on average, has effects over a 5-year period. Additional analysis details and results are presented in the **Supplementary Materials**.

## Discussion

In this economic analysis, we quantified both the cost effectiveness and budget impact of a Fitbit intervention relative to usual care in an insufficiently active, middle-aged population of U.S. adults. Over a 15-year time horizon, the CEA demonstrated that the Fitbit intervention would be cost saving and result in more QALYs than if participants only received usual care. When assessed over a shorter time horizon from a hypothetical Commercial health plan perspective, the BIA results showed the Fitbit intervention would save $6-8 million dollars over a 2-to 5-year period for a cohort of plan participants. The results were most sensitive to assumptions about short-term healthcare cost impacts but the cost-saving conclusions remained consistent across all scenario analyses that explored model and parameter uncertainty.

The overall findings are consistent with prior health economic models of interventions that aim to increase physical activity through the use of consumer wearable activity trackers, particularly those that leverage pedometer-based interventions. A systematic review and modeling analysis by Gc et. al. reported that brief interventions leveraging pedometers as a motivational tool (with or without additional exercise counseling) were likely to be cost-saving at a population level.^39,97^ Specifically, a program encouraging the use of pedometers was more effective and less costly when compared to usual care in the general Australian population; pairing a pedometer with a step-based exercise prescription goal was similarly dominant when compared to exercise prescription alone for those older than age 65 in New Zealand.^32,98^ In Belgium, a multi-strategy health promotion campaign that incorporated pedometer use was also dominant when compared to no intervention among a general adult population.^99^

Our findings of short-term cost reductions from a fully remotely-delivered intervention have also been demonstrated among adults receiving primary care in the U.K. Anokye et. al.^30^ found that postal delivery of a pedometer intervention had a 50% chance of being cost-effective after 1 year of the intervention compared to control, and is dominant in the long-term when compared to both usual care and a version of the pedometer intervention that included nurse consultations. In particular, short-term cost savings in the postal pedometer group in the first year were due to lower health services use.^30^ While the modeled Fitbit intervention differs slightly in its mode of implementation (i.e. not delivered within primary care practice), Fitbit’s robust tracking of physical activity metrics, including step counts and active minutes, as well as goal setting and feedback features compares favorably to pedometer interventions that may be delivered during a brief healthcare encounter.

The strengths of our research include its robust model design and conservative approach to model parameters and assumptions. The model concept and structure were informed by, and are generally consistent with, numerous other economic analyses of physical activity interventions.^30–39^ One substantial enhancement over older analyses was the use of mean daily step count improvements from RCTs of Fitbit-based interventions and longitudinal health outcomes from over 6,000 Fitbit users with 4 years of median follow-up to estimate rates of incident disease. When calculating step count improvement, we used the most conservative approach to bias results against the Fitbit intervention. Finally, our PSA was extensive, including over 40 key model parameters, strengthening the confidence and robustness of our findings.

Despite its strengths, our study has important limitations. First, although there is abundant literature documenting short-term cost savings associated with improved physical activity,^47–52^ most of those studies were conducted in older patients which limits the generalizability of those results to our middle-aged cohort. We attempted to minimize the effect of this data issue by making conservative assumptions, thus reducing the cost offset of the Fitbit intervention due to potential differences in population age and intervention efficacy. In addition, our PSA allowed for short-term costs to be greater in the Fitbit intervention than control. Finally, even without short-term cost savings, our analysis suggests that a Fitbit intervention is likely to still be cost-saving compared to usual care.

Next, although the Fitbit intervention has many important elements other than simply providing daily step count information (e.g., app-based goal setting and personalized feedback that may substitute for similar elements that would have incurred additional costs if provided by clinical staff, physical therapist, etc.), our analysis does not use *de novo* data in the population described. Rather, the analysis relies on RCTs that may not exactly replicate the intervention as described or the modeled population, nor have sufficient follow-up to inform the longer-term impact on physical activity levels beyond the duration of the intervention. This issue was addressed with sensitivity analyses that varied the intervention efficacy (in terms of daily steps) and durability (time to return to baseline step counts).

Finally, we acknowledge limitations in how our model structure captures the benefits of increased levels of physical activity. Estimates of disease incidence stratified by daily step counts in our model were based on reported associations rather than causal estimates. While an RCT would be the optimal approach to prove causality; in the absence of such data, we leveraged detailed disease incidence estimates from a robust longitudinal cohort study that provided multiple years of objectively tracked levels of physical activity, as measured by Fitbit devices. These estimates are likely a better reflection of physical activity levels under real-world conditions as opposed to prior estimates that relied on self-reports or short-term accelerometry measurements. We additionally varied these disease incidence estimates broadly in multi-way sensitivity analyses. Across these analyses, the Fitbit intervention remained very likely to be cost-saving compared to usual care. In addition, since the present analysis characterized the benefits of physical activity only through reduced disease incidence among the ‘Well’ population, it is possible that the overall magnitude of benefits could be even larger if improvements in chronic condition management were also included.

Our research strengthens support for implementing wearable-based interventions to improve population health through increasing physical activity among insufficiently active adults. We illustrate the health economic value of deploying a relatively low-cost wearable-based intervention, as it can be potentially cost-saving through lower healthcare utilization and disease-related costs, even if the resulting improvements in clinical outcomes are modest. Taken together, this offers compelling support for healthcare payers to consider including wearable-based physical activity interventions as part of a comprehensive portfolio of preventive health offerings for their insured populations.

## Data Availability

All data produced in the present study are available upon reasonable request to the authors.

## Acknowledgements

We gratefully acknowledge Amy McDonough and the Strategic Health Solutions team for supporting this study, and Jeffrey Annis (Vanderbilt Institute for Clinical and Translational Research) for his assistance with parameter estimates used in our modeling analyses.

## SUPPLEMENTARY MATERIALS

### Supplementary Text

#### Calculation of step improvement for Fitbit

The increase in daily step count associated with the Fitbit intervention was estimated from 8 RCTs that compared a Fitbit-based intervention to a control group without the device.^55–62^ The mean step count difference between the two comparators was calculated using two approaches: 1) a “mean change” or difference in difference approach that accounts for disparities in the baseline step counts for the Fitbit and control groups; and 2) a “post-intervention” approach that assumes baseline step counts are not significantly different between the two groups and only considers the difference in the final step count estimates. In their meta-analysis of Fitbit RCTs, Ringeval and colleagues (2020)^23^ used a mix of these two approaches in their calculations. We preferred consistency and chose the “mean change” approach for our estimation as it yielded a more conservative result (mean difference of +1,287 steps) compared to the “post-intervention” approach (+1,677 steps).

#### Evaluation of model structural uncertainty

As recommended in good research practice guidelines,^41,42,44^ model structural uncertainty should be evaluated. The first method employed to meet this recommendation was to substitute a “tiered steps approach”’ for the “mean steps approach” used as the model default. The latter uses the simple weighted average daily step improvement quantified from RCTs using a Fitbit device.^55–62^ The tiered steps approach assumes a more complex (and realistic) structure in which the mean daily steps are stratified into 3 tiers at baseline: 1) 5,500 daily steps (25% of cohort); 2) 6,500 daily steps (50%); and 3) 7,500 daily steps (25%). As with the base case, for usual care, there is no improvement in mean daily steps. For the Fitbit intervention for the 3 groups, there is improvement of 0 steps in tier 1, 1,200 daily steps in tier 2, and 2,700 steps in tier 3. The distribution and daily step count improvement are arbitrary but net out to similar baseline and maximum improvement step counts as with the base case—6,500 and 1,275, respectively. This attempt to instill heterogeneity into the population and daily step count model structure appears to have minimal impact on the results.

A second approach for examining structural uncertainty pertains to the types of resource use and costs that might be considered in the initial 2-year period. In the base case, short-term costs consisted solely of adjusted estimates from a U.S. economic analysis of the Silver Sneakers program.^52^ It was uncertain whether inclusion of health state costs (‘Well’ and incident diseases) in the initial 2-year period would represent a more thorough accounting of health costs or the degree to which some costs would be double-counted. Therefore, a modification to the model programming was made to allow for health state cost inclusion in all model cycles. As expected, given that the Fitbit intervention leads to reductions in incident disease, inclusion of those costs in the first two years led to a modest increase in cost savings. While the conclusions of the analysis remained unchanged with this assessment of model uncertainty, we maintain that the conservative method (exclusion of health state costs in the initial 2-year period) remains the desired base case approach.

#### Exploratory analysis of an older, insufficiently active population

The base case model and analysis was designed to estimate the impact of the Fitbit intervention on physical activity in a middle-aged cohort (starting age = 50) in terms of an increase in the number of daily steps and reductions in medical resource use, costs, and incident disease. Few economic analyses have evaluated physical activity interventions in older adults (age ≥65) in terms of daily steps and none, to our knowledge, have linked steps to a robust source of incident disease data which remains a gap warranting future research. However, a recent economic analysis by Deidda and colleagues (2022)^31^ in older adults did provide sufficient data to inform an exploratory analysis of the cost effectiveness of a Fitbit intervention in a Medicare population.

The Deidda study examined the cost-effectiveness of physical activity interventions (an exercise referral scheme [ERS] with or without self-management strategies [SMS]) using a long-term economic model extending the analysis of the SITLESS randomized controlled trial in older adults. The model used two physical activity health states (‘active’ and ‘inactive’), corresponding to maintaining (or not) at least 150 minutes of moderate to vigorous aerobic physical activity, and seven health states representing conditions associated with an inactive lifestyle in an older population (e.g., Type 2 diabetes, Alzheimer’s disease, falls, etc.). This approach differs from our model concept which uses daily step counts and chronic diseases more relevant to a middle-aged population. Therefore, some modification to both model structure and parameter estimates was required for our exploratory analysis. Furthermore, given that Medicare is the predominant U.S. payer for a population aged 65+, the time horizon for the cost-effectiveness analysis was increased from 15 years to a lifetime horizon.

### Population

The target population of interest for the exploratory analysis was older adults with insufficient physical activity, aged ≥65 (starting age=65), insured by Medicare. The age and sex distribution was based on 2022 data from the U.S. Census Bureau.^53^ The proportion with insufficient physical activity in this population was informed by a systematic review of 23 published reports of the prevalence of sedentary behavior in older adults.^100^ Population details are shown in **Supplementary Table 4**.

### Comparators

Unlike our base case analysis, Deidda compared Usual Care to physical activity interventions that included an exercise referral scheme. Such schemes have a mixed history with some economic analyses suggesting unfavorable cost effectiveness,^32,38^ favorable cost effectiveness,^35^ or as Deidda reported, potential cost savings. In their analysis, Deidda and colleagues used data from the SITLESS RCT to generate probabilities of moving between the two physical activity health states for each of the modeled comparators. A different set of transition probabilities was used for the first year in the analysis than all subsequent years. To estimate the incidence of acute (i.e., falls) and chronic conditions (e.g., Alzheimer’s disease) related to insufficient physical activity in the older population over time, the transition probabilities and published incidence data (stratified by activity level) were utilized. Unit costs, utilities, and mortality rates were then applied to the acute/chronic health states to calculate total costs and QALYs (**Supplementary Table 4**).

Our model structure was modified to better accommodate the approach used by Deidda and colleagues. First, a review of U.S. costs, utilities, disutilities, and mortality for the health state conditions in the Deidda model was conducted. Based on the findings, the health states included in the exploratory analysis were: 1) Alzheimer’s disease/dementia; 2) Type 2 diabetes; 3) cardiovascular disease; and 4) falls. Second, the ERS transition probabilities were adjusted so that the annual proportion of patients in the active and inactive health states for Fitbit were equivalent to Usual Care at 5 years and beyond (a similar durability assumption to our base case) rather than maintaining an advantage in physical activity relative to Usual Care as was the case for ERS in the Deidda study. Third, the health state distributions were used in conjunction with the disease incidence data to calculate a weighted average incidence for each condition for each comparator. These incidence estimates were then applied recursively over time to the proportion alive in each annual model cycle to calculate the proportion in each of the health states. Finally, unit costs, utilities/disutilities, and mortality rates were applied to each health state to calculate total costs and QALYs.

### Costs

As in the base case analysis, the costs in the exploratory model include those related to the Fitbit intervention (incremental to usual care which is assumed to have no cost), short-term healthcare costs (2 years), and health state costs (‘Well’ and incident conditions incremental to the ‘Well’ health state). Unit costs for the model were derived primarily from published sources.^52,64,68,70,101–103^ When applicable, published costs were inflated to 2023 USD using the Medical Care component of the U.S. CPI (**Supplementary Table 4**).^72^

In the exploratory analyses, we assumed a greater potential for some short-term cost reduction given the similarity in our population to the Medicare Advantage enrollees participating in the Silver Sneakers^®^ (SS) healthy aging program.^52^ Because the SS intervention is not an exact match to those in our model, we maintained the efficacy reduction factor from the base case analysis (assuming the SS program has a greater impact on resource use/cost). Longer-term costs associated with the model health states (‘Well’ and incident acute and chronic conditions) were quantified for both comparators in a similar fashion to the base case analysis.

### Mortality

The annual probability of death for participants in the ‘Well’ health state was based on age- and sex-adjusted all-cause mortality data from U.S. life tables.^73^ Published hazard ratios^77,81,104,105^ describing the excess mortality associated with each of the health conditions of interest were used to adjust background all-cause mortality and the annual probability of death for those diseases. Falls were assumed to be an acute event that could occur in any cycle with a conditional probability of death for any fall event (**Supplementary Table 4**).^103^

### Utilities

Utilities for the ‘Well’ health state (by age strata) were based on a nationally representative catalog of EuroQol-5 Dimension (EQ-5D) utility scores.^84^ For the acute/chronic conditions in the model, we identified published disutilities^106–108^ (or utilities that were subsequently used to derive disutilities using information in Sullivan, et al. 2006^84^) that were applied to the ‘Well’ health state utility to account for the worse quality of life for those conditions.

**Supplementary Table 1.** Annual Incidence Rates for Chronic Diseases in the Model^a^.

| Chronic Disease | Year 0 | Year 1 | Year 2 | Year 3 | Year 4 | Year 5 | Year 6 | Year 7 | Year 8 | Year 9 | Year 10 | Year 11 |
| --- | --- | --- | --- | --- | --- | --- | --- | --- | --- | --- | --- | --- |
| **Usual Care** |  |  |  |  |  |  |  |  |  |  |  |  |
| Obesity | 0.00% | 1.01% | 1.02% | 1.03% | 2.10% | 2.14% | 2.85% | 2.93% | 3.23% | 3.55% | 3.90% | 4.29% |
| Type 2 diabetes | 0.00% | 0.26% | 0.26% | 0.26% | 0.51% | 0.52% | 0.64% | 0.65% | 0.71% | 0.78% | 0.86% | 0.95% |
| Hypertension | 0.00% | 1.34% | 1.36% | 1.38% | 3.28% | 3.39% | 4.02% | 4.19% | 4.61% | 5.07% | 5.58% | 6.14% |
| GERD | 0.00% | 1.75% | 1.78% | 1.81% | 3.16% | 3.27% | 4.50% | 4.72% | 5.19% | 5.71% | 6.28% | 6.91% |
| MDD | 0.00% | 1.40% | 1.42% | 1.44% | 2.87% | 2.95% | 3.84% | 3.99% | 4.39% | 4.83% | 5.31% | 5.84% |
| Sleep apnea | 0.00% | 0.92% | 0.93% | 0.94% | 1.77% | 1.80% | 1.97% | 2.01% | 2.21% | 2.43% | 2.68% | 2.95% |
| **Fitbit Intervention** |  |  |  |  |  |  |  |  |  |  |  |  |
| Obesity | 0.00% | 0.90% | 0.93% | 0.96% | 2.05% | Equivalent to Usual Care |  |  |  |  |  |  |
| Type 2 diabetes | 0.00% | 0.24% | 0.24% | 0.25% | 0.50% |  |  |  |  |  |  |  |
| Hypertension | 0.00% | 1.23% | 1.26% | 1.31% | 3.19% |  |  |  |  |  |  |  |
| GERD | 0.00% | 1.58% | 1.65% | 1.73% | 3.12% |  |  |  |  |  |  |  |
| MDD | 0.00% | 1.25% | 1.29% | 1.36% | 2.76% |  |  |  |  |  |  |  |
| Sleep apnea | 0.00% | 0.79% | 0.82% | 0.87% | 1.69% |  |  |  |  |  |  |  |

Sources: STEPS—Amorim 2019;<sup>55</sup> Ashe 2015;<sup>56</sup> Cadmus-Bertram 2019;<sup>57</sup> Duscha 2018;<sup>58</sup> Katz 2018;<sup>59</sup> Li 2018;<sup>60</sup> Paxton 2018;<sup>61</sup> Van Blarigan 2019;<sup>62</sup> INCIDENCE AT STEP COUNT—Master 2022<sup>13</sup>
Abbreviations: GERD, gastroesophageal reflux disease; MDD, major depressive disorder
<sup>a</sup>Incidence estimates from the All of Us study concluded at Year 7; therefore an annual growth rate factor of 10% was applied in Years 8-11 to simulate the increasing incidence over time observed in the All of Us Study; starting at Year 11, incidence rates were held constant for the remainder of the analysis

**Supplementary Table 2.** Parameters Used in Probabilistic Sensitivity Analyses (Cost Effectiveness Only)

| Parameter | Value | SE | Distribution | Source Informing SE<sup>b</sup> |
| --- | --- | --- | --- | --- |
| **Intervention Details** |  |  |  |  |
| Baseline number of daily steps | 6,500 | 125 | Normal | Ringeval 2020<sup>23</sup> |
| Mean peak change in daily steps for Fitbit intervention | 1,287 | 125 | Normal | Ringeval 2020<sup>23</sup> |
| Durability of Fitbit intervention (years)<sup>a</sup> | 5 | 0.05 | Normal | Chaudhry 2020;<sup>19</sup> Gasana 2023,<sup>63</sup> assumption |
| **Clinical Details** |  |  |  |  |
| % of diabetes with comorbid obesity<sup>a</sup> | 50% | 10% | Beta | Assumption |
| % of diabetes with comorbid hypertension<sup>a</sup> | 50% | 10% | Beta | Assumption |
| Excess all-cause mortality hazard ratios |  |  |  |  |
| Obesity (age 35-49) | 2.40 | 0.08 | Lognormal | Ward 2022<sup>83</sup> |
| Obesity (age 50-69) | 1.90 | 0.05 | Lognormal | Ward 2022<sup>83</sup> |
| Obesity (age 70+) | 1.50 | 0.03 | Lognormal | Ward 2022<sup>83</sup> |
| Type 2 diabetes | 1.95 | 0.16 | Lognormal | Preis 2009<sup>81</sup> |
| Hypertension | 1.41 | 0.03 | Lognormal | Aune 2021<sup>74</sup> |
| Major depressive disorder | 1.70 | 0.13 | Lognormal | Pratt 2016;<sup>80</sup> Gilman 2017<sup>76</sup> |
| Sleep apnea | 1.40 | 0.20 | Lognormal | Lin 2023<sup>78</sup> |
| Annual incidence growth rate factor after 7 years | 10% | 1% | Beta | Assumption |
| **Healthcare Cost Details<sup>c</sup>** |  |  |  |  |
| Annual short-term healthcare costs (Years 1 and 2) |  |  |  |  |
| Usual care (per participant) | \$8,687 | \$869 | Gamma | Gc 2018;<sup>39</sup> assumption |
| Fitbit intervention (per participant) | \$7,268 | \$727 | Gamma | Gc 2018;<sup>39</sup> assumption |
| Age-based adjustment factor | 50% | 10.8% | Beta | Assumption |
| Efficacy-based adjustment factor | 25% | 10.8% | Beta | Assumption |
| Annual health state costs |  |  |  |  |
| Age 40-64 |  |  |  |  |
| Well | \$4,720 | \$472 | Gamma | Gc 2018;<sup>39</sup> assumption |
| Obesity | \$1,093 | \$109 | Gamma | Gc 2018;<sup>39</sup> assumption |
| Type 2 diabetes | \$9,673 | \$967 | Gamma | Gc 2018;<sup>39</sup> assumption |
| Hypertension | \$3,001 | \$300 | Gamma | Gc 2018;<sup>39</sup> assumption |
| Diabetes + obesity | \$14,179 | \$1,418 | Gamma | Gc 2018;<sup>39</sup> assumption |
| Diabetes + hypertension | \$12,072 | \$1,207 | Gamma | Gc 2018;<sup>39</sup> assumption |
| GERD | \$7,341 | \$734 | Gamma | Gc 2018;<sup>39</sup> assumption |
| Major depressive disorder | \$6,886 | \$689 | Gamma | Gc 2018;<sup>39</sup> assumption |
| Sleep apnea | \$9,222 | \$922 | Gamma | Gc 2018;<sup>39</sup> assumption |
| Age ≥65 |  |  |  |  |
| Well | \$6,554 | \$655 | Gamma | Gc 2018;<sup>39</sup> assumption |
| Obesity | \$1,327 | \$133 | Gamma | Gc 2018;<sup>39</sup> assumption |
| Type 2 diabetes | \$9,882 | \$988 | Gamma | Gc 2018;<sup>39</sup> assumption |
| Hypertension | \$2,853 | \$285 | Gamma | Gc 2018;<sup>39</sup> assumption |
| Diabetes + obesity | \$11,561 | \$1,156 | Gamma | Gc 2018;<sup>39</sup> assumption |
| Diabetes + hypertension | \$9,713 | \$971 | Gamma | Gc 2018;<sup>39</sup> assumption |
| GERD<sup>c</sup> | \$4,918 | \$492 | Gamma | Gc 2018;<sup>39</sup> assumption |
| Major depressive disorder | \$4,614 | \$461 | Gamma | Gc 2018;<sup>39</sup> assumption |
| Sleep apnea | \$11,538 | \$1,154 | Gamma | Gc 2018;<sup>39</sup> assumption |
| **Disutility Details<sup>d</sup>** |  |  |  |  |
| Obesity | -0.127 | -0.0009 | Beta | Sullivan 2006<sup>84</sup> |
| Type 2 diabetes | -0.071 | -0.0004 | Beta | Sullivan 2006<sup>84</sup> |
| Hypertension | -0.057 | -0.0005 | Beta | Sullivan 2006<sup>84</sup> |
| GERD | -0.058 | -0.0005 | Beta | Sullivan 2006<sup>84</sup> |
| Major depressive disorder | -0.156 | -0.0036 | Beta | Hanmer 2016;<sup>85</sup> Sullivan 2006<sup>84</sup> |
| Sleep apnea | -0.075 | -0.0126 | Beta | Hanmer 2016;<sup>85</sup> Walia 2017<sup>90</sup> |

Abbreviations: GERD, gastroesophageal reflux disease; SE, standard error
<sup>a</sup>Some parameters were truncated to ensure realistic values—Fitbit durability forced to be in the 2–6 year range; diabetes comorbidity percentages ≤50%; <sup>b</sup>When assumptions were required for a standard error estimate, 95% confidence intervals were approximated to ensure that ranges over which the parameter would vary were reasonable; <sup>c</sup>In most cases, published costs were not reported with a measure of uncertainty; therefore, a standard error of 10% was assumed as was the approach in Gc 2018<sup>39</sup> (an economic analysis of physical activity interventions); <sup>d</sup>Disutilities may be expressed as negative numbers but a beta distribution is limited to positive numbers; therefore, the absolute values of the disutility and standard errors were used and then the sampled estimate used in the model was converted to a negative number

**Supplementary Figure 1.**
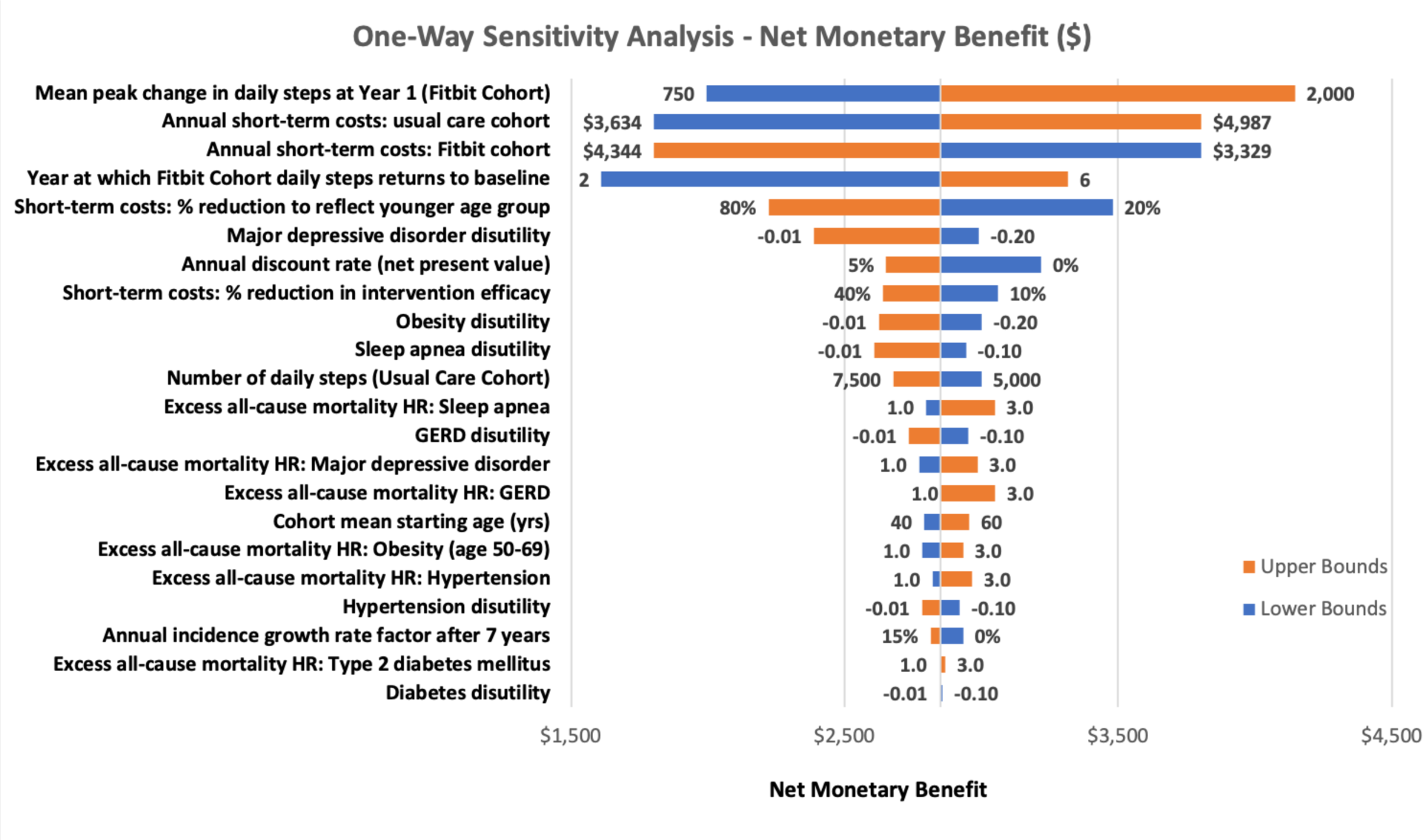
One-way sensitivity analysis tornado diagram of net monetary benefit (at a willingness to pay threshold of $150,000/QALY gained) in the cost-effectiveness analysis. Lower and upper bound parameter estimates for each variable are listed within the diagram. Abbreviations: GERD, gastroesophageal reflux disease; HR, hazard ratio; yrs, years

**Supplementary Table 3.** Additional Scenario Analysis Results.

|  | Costs |  |  | QALYs |  |  | ICER or PMPM |
| --- | --- | --- | --- | --- | --- | --- | --- |
|  | Usual Care | Fitbit | Incremental | Usual Care | Fitbit | Incremental |  |
| **Base Case** |  |  |  |  |  |  |  |
| Cost effectiveness<sup>a</sup> | \$89,541 | \$88,284 | -\$1,257 | 9.031 | 9.042 | 0.011 | Fitbit dominant<sup>b</sup> |
| Budget impact<sup>a</sup> | \$220,166,612 | \$211,617,969 | -\$8,548,644 | NA | NA | NA | -\$0.14 |
| **Structural Uncertainty** |  |  |  |  |  |  |  |
| **Tiered steps approach<sup>c</sup>** |  |  |  |  |  |  |  |
| Cost effectiveness | \$89,539 | \$88,242 | -\$1,297 | 9.032 | 9.041 | 0.009 | Fitbit dominant<sup>b</sup> |
| Budget impact | \$220,169,841 | \$211,713,811 | -\$8,456,030 | NA | NA | NA | -\$0.14 |
| **Inclusion of health state costs in Years 1 & 2** |  |  |  |  |  |  |  |
| Cost effectiveness | \$99,888 | \$98,522 | -\$1,367 | 9.031 | 9.042 | 0.011 | Fitbit dominant<sup>b</sup> |
| Budget impact | \$304,209,839 | \$294,766,789 | -\$9,443,050 | NA | NA | NA | -\$0.16 |
| **Scenarios** |  |  |  |  |  |  |  |
| **Use of post-intervention steps estimate** |  |  |  |  |  |  |  |
| Cost effectiveness | \$89,541 | \$88,173 | -\$1,368 | 9.031 | 9.045 | 0.014 | Fitbit dominant<sup>b</sup> |
| Budget impact | \$220,166,612 | \$211,061,720 | -\$9,104,892 | NA | NA | NA | -\$0.15 |
| **Reduced population eligibility and adoption rate<sup>d</sup>** |  |  |  |  |  |  |  |
| Budget impact | \$110,083,306 | \$105,808,984 | -\$4,274,322 | NA | NA | NA | -\$0.07 |
| **Increased population eligibility and adoption rate<sup>d</sup>** |  |  |  |  |  |  |  |
| Budget impact | \$330,249,918 | \$317,426,953 | -\$12,822,965 | NA | NA | NA | -\$0.21 |
| **Reduced cumulative incidence of chronic disease<sup>e</sup>** |  |  |  |  |  |  |  |
|  | Usual Care | Fitbit | Incremental | Usual Care | Fitbit | Incremental |  |
| Cost effectiveness | \$85,736 | \$84,540 | -\$1,196 | 9.105 | 9.114 | 0.009 | Fitbit dominant<sup>b</sup> |
| Budget impact | \$212,916,291 | \$204,778,351 | -\$8,137,940 | NA | NA | NA | -\$0.14 |
| **Increased cumulative incidence of chronic disease<sup>e</sup>** |  |  |  |  |  |  |  |
| Cost effectiveness | \$92,582 | \$91,280 | -\$1,302 | 8.972 | 8.984 | 0.012 | Fitbit dominant<sup>b</sup> |
| Budget impact | \$227,009,258 | \$218,097,985 | -\$8,911,272 | NA | NA | NA | -\$0.15 |
| **Lifetime horizon** |  |  |  |  |  |  |  |
| Cost effectiveness | \$162,005 | \$160,710 | -\$1,295 | 13.463 | 13.476 | 0.013 | Fitbit dominant<sup>b</sup> |
| **Health state costs only<sup>f</sup>** |  |  |  |  |  |  |  |
| Cost effectiveness | \$91,328 | \$91,010 | -\$318 | 9.031 | 9.042 | 0.011 | Fitbit dominant<sup>b</sup> |
| Budget impact | \$234,713,839 | \$233,784,789 | -\$929,050 | NA | NA | NA | -\$0.02 |

Abbreviations: ICER, incremental cost-effectiveness ratio; PMPM, per-member per-month; QALY, quality-adjusted life-year
<sup>a</sup>15-year time horizon for CEA, 5 year horizon for BIA; <sup>b</sup>“Dominant” indicates that Fitbit is less costly and more effective than Usual Care; <sup>c</sup>The tiered approach uses the following 3 strata (“tiers”) of step counts for Usual Care (25% with 5,500 mean daily steps; 50% with 6,500 mean daily steps; and 25% with 7,500 mean daily steps) none of whom have increased mean daily step counts over time; the 3 strata for the Fitbit intervention are the same at baseline but have maximum improvements of 0 steps (strata 1), 1,200 steps (strata 2), and 2,700 steps (strata 3); <sup>d</sup>The base case adoption rate estimate (5%) was decreased and increased to 2.5% and 7.5%, respectively; <sup>e</sup>Chronic disease cumulative incidence estimates were set to the lower and upper limits of the linear function obtained from the *All of Us* researchers; <sup>f</sup>Short-term healthcare costs in Years 1 and 2 were eliminated and health state costs—‘Well’ and incident diseases—added to the first 2 years

**Supplementary Table 4.** Exploratory Analysis Model Parameter Estimates and Sources.

| Parameter | Value | Sources |
| --- | --- | --- |
| **Population Details** |  |  |
| Health plan size (# members)<sup>a</sup> | 1,000,000 | Assumption |
| % of plan aged 65+<sup>a</sup> | 100% | US Census Bureau 2023<sup>53</sup> |
| % of cohort that is insufficiently active<sup>a</sup> | 67% | Harvey 2013<sup>100</sup> |
| % eligible and willing to adopt Fitbit intervention<sup>a</sup> | 5% | Assumption |
| Cohort mean starting age (years) | 65 | Assumption |
| % female | 55% | US Census Bureau 2023<sup>53</sup> |
| **Intervention Details** |  |  |
| Usual care cost | \$0 | Assumption |
| Fitbit intervention cost | \$250 | Retail Fitbit pricing<sup>b</sup>; assumptions |
| Durability of Fitbit intervention (years)<sup>c</sup> | 5 | Deidda 2022,<sup>31</sup> assumptions |
| **Clinical Details** |  |  |
| Background all-cause mortality | Age-based | Arias 2023<sup>73</sup> |
| Annual acute condition/chronic disease incidence |  |  |
| Type 2 diabetes (active) | 0.0034 | InterAct Consortium 2012<sup>109</sup> in Deidda 2022<sup>31</sup> |
| Type 2 diabetes (inactive) | 0.0049 | InterAct Consortium 2012<sup>109</sup> in Deidda 2022<sup>31</sup> |
| Alzheimer's disease/dementia (active) | 0.0125 | Abbott 2004;<sup>110</sup> Podewils 2005;<sup>111</sup> Larsson 2018<sup>112</sup> in Deidda 2022<sup>31</sup> |
| Alzheimer's disease/dementia (inactive) | 0.0167 | Abbott 2004;<sup>110</sup> Podewils 2005;<sup>111</sup> Larsson 2018<sup>112</sup> in Deidda 2022<sup>31</sup> |
| Cardiovascular disease (active) | 0.0255 | Jefferis 2019;<sup>113</sup> Lacey 2015,<sup>114</sup> Patel 2020<sup>115</sup> in Deidda 2022<sup>31</sup> |
| Cardiovascular disease (inactive) | 0.0410 | Jefferis 2019;<sup>113</sup> Lacey 2015,<sup>114</sup> Patel 2020<sup>115</sup> in Deidda 2022<sup>31</sup> |
| Falls (active) | 0.0776 | Buchner 2017<sup>116</sup> in Deidda 2022<sup>31</sup> |
| Falls (inactive) | 0.1085 | Buchner 2017<sup>116</sup> in Deidda 2022<sup>31</sup> |
| Excess all-cause mortality hazard ratios or %<sup>c</sup> |  |  |
| Type 2 diabetes | 1.95 | Preis 2009;<sup>81</sup> Li 2019<sup>77</sup> |
| Alzheimer's disease/dementia | 2.00 | Lanctot 2024<sup>104</sup> |
| Cardiovascular disease | 1.40 | Shaked 2022<sup>105</sup> |
| Falls | 0.75% | Burns 2016<sup>103</sup> |
| **Healthcare Cost Details<sup>d</sup>** |  |  |
| Annual short-term healthcare costs (Years 1 and 2) |  |  |
| Usual care (per participant) | \$8,687 | Teigland 2022<sup>52</sup> |
| Fitbit intervention (per participant) | \$7,268 | Teigland 2022;<sup>52</sup> assumption |
| Efficacy-based adjustment factor | 25% | Assumption |
| Annual health state costs<sup>e</sup> |  |  |
| Well | \$6,554 | Leung 2017;<sup>68</sup> Wang 2017<sup>70</sup> |
| Type 2 diabetes | \$9,882 | ADA 2018;<sup>64</sup> Leung 2017;<sup>68</sup> Wang 2017<sup>70</sup> |
| Alzheimer's disease/dementia | \$31,193 | Alzheimer's Association 2023<sup>101</sup> |
| Cardiovascular disease | \$5,066 | Birger 2021<sup>102</sup> |
| Falls | \$12,650 | Burns 2016<sup>103</sup> |
| **Utility/Disutility Details<sup>f</sup>** |  |  |
| Well |  |  |
| Age 60-69 | 0.823 | Sullivan 2006<sup>84</sup> |
| Age 70-79 | 0.790 | Sullivan 2006<sup>84</sup> |
| Age ≥80 | 0.736 | Sullivan 2006<sup>84</sup> |
| Type 2 diabetes | -0.071 | Sullivan 2006<sup>84</sup> |
| Alzheimer's disease/dementia | -0.180 | Vandepitte 2020<sup>108</sup> in Deidda 2022<sup>31</sup> |
| Cardiovascular disease | -0.060 | Lacey 2003<sup>107</sup> in Deidda 2022<sup>31</sup> |
| Falls | -0.160 | Bjerk 2019<sup>106</sup> in Deidda 2022<sup>31</sup> |
| **Miscellaneous Analysis Details** |  |  |
| Budget impact time horizon (years) | 2 and 5 | Assumption |
| Cost-effectiveness time horizon (years) | Lifetime | Assumption |
| Annual discount rate (costs, QALYs)<sup>g</sup> | 3% | Lipscomb 1996;<sup>93</sup> Weinstein 1996<sup>94</sup> |

Abbreviations: QALY, quality-adjusted life-year
<sup>a</sup>Budget impact analysis only; <sup>b</sup>The cost of the Fitbit intervention is estimated based on the retail price of a Charge 6 device, in addition to a 12-month subscription to Fitbit Premium (which provides goal setting, feedback, and access to a library of workout content and additional detailed metrics in-app), as well as additional intervention deployment costs; <sup>c</sup>Unlike the base case analysis, the exploratory analysis does not use daily step count as an efficacy measure but rather uses the transition probabilities in Deidda 2022<sup>31</sup> as an efficacy measure and for calculation of health condition incidence; using adjusted transition probabilities and incidence estimates from the Deidda study, annual incidence rates converge between the Usual Care and Fitbit groups by 5 years; <sup>d</sup>Costs from published literature not reported in 2023 USD were inflated using the Medical Care consumer price index; <sup>e</sup>Incident disease costs are incremental to the 'Well' health state cost; <sup>f</sup>Disutilities were used for incident diseases to avoid the counterintuitive result in which the utility of an incident disease health state is higher (more favorable) than the 'Well' state which could occur at older ages; <sup>g</sup>Cost-effectiveness analysis only

**Supplementary Table 5.** Exploratory Economic Analysis Results.

|  | Usual Care | Fitbit | Incremental |
| --- | --- | --- | --- |
| **Cost Effectiveness<sup>a</sup>** |  |  |  |
| Total costs | \$148,776 | \$146,798 | -\$1,977 |
| Total LYs | 12.131 | 12.135 | 0.004 |
| Total QALYs | 9.921 | 9.923 | 0.002 |
| ICER (cost/LY gained) |  |  | Fitbit dominates UC<sup>c</sup> |
| ICER (cost/QALY gained) |  |  | Fitbit dominates UC<sup>c</sup> |
| **Budget Impact<sup>b</sup>** |  |  |  |
| 2-year total costs | \$582,029,000 | \$510,724,250 | -\$62,929,750 |
| 2-year incremental budget impact (PMPM) | | | -\$2.62 |
| 5-year total costs | \$1,575,338,659 | \$1,509,713,110 | -\$65,625,550 |
| 5-year incremental budget impact (PMPM) | | | -\$1.09 |

Abbreviations: ICER, incremental cost-effectiveness ratio; LY, life-year; PMPM, per-member per-month; QALY, quality-adjusted life-year; UC, usual care
<sup>a</sup>Expected costs and outcomes per participant quantified over a lifetime horizon, discounted at 3% annually; <sup>b</sup>Costs for a cohort of 33,500 participants over both a 2-year and 5-year time horizon (undiscounted); <sup>c</sup>“Dominates” indicates that Fitbit is less costly and more effective than UC

**Supplementary Figure 2.**
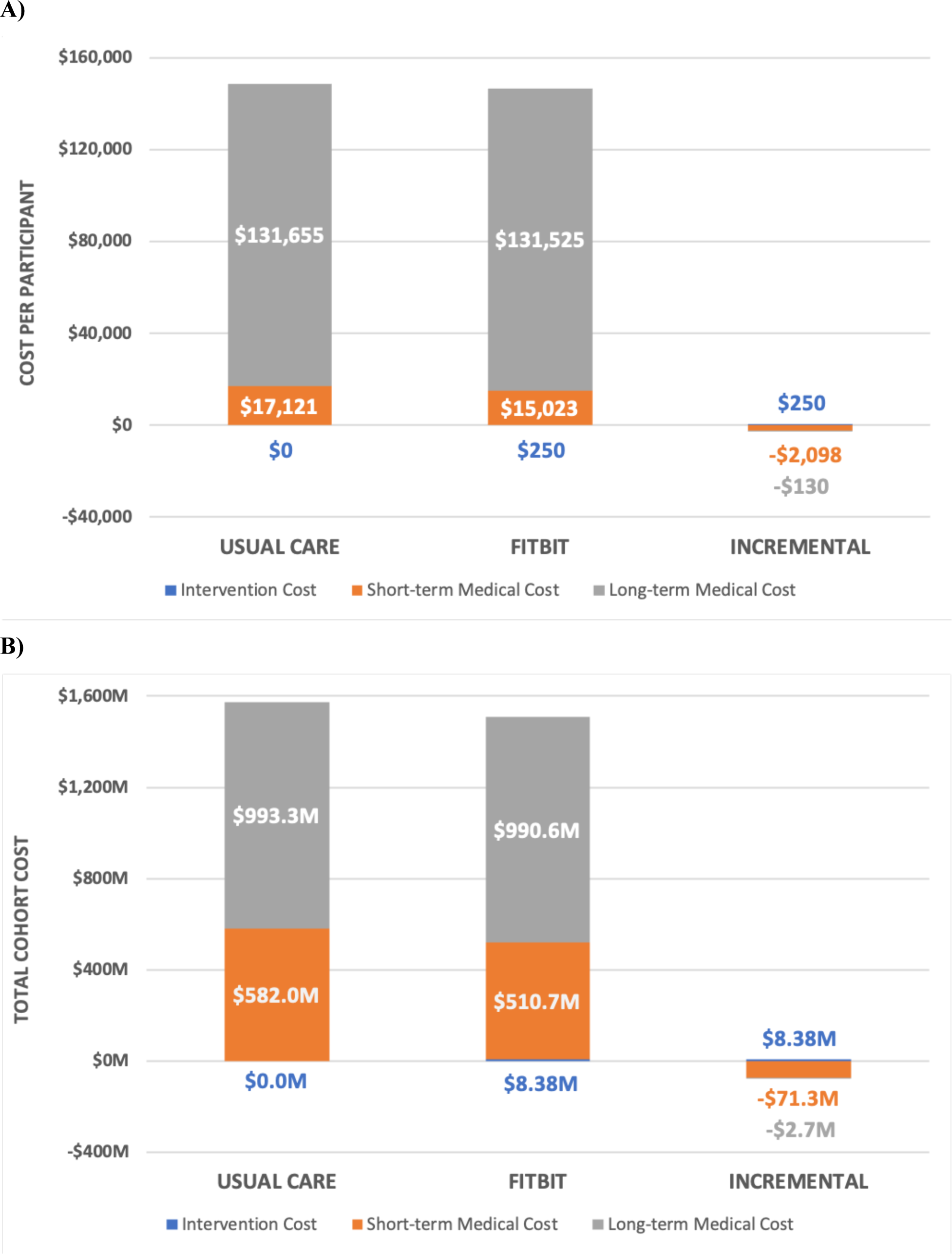
Stratified cost results of the exploratory A) cost-effectiveness and B) budget impact analyses. The time horizons of the analyses are lifetime and 5 years, respectively. The majority of the cost savings is associated with reductions in healthcare resource use in the first two years after initiation of the interventions.

